# Alzheimer’s Disease cerebrospinal fluid Biomarkers and kidney function in normal and cognitively impaired older adults

**DOI:** 10.1101/2023.11.01.23297910

**Authors:** Ihab Hajjar, Reem Neal, Zhiyi Yang, James J. Lah

## Abstract

**Importance:** Recent Alzheimer’s disease (AD) clinical trials have used Cerebrospinal fluid (CSF) biomarker levels for screening and enrollment. Preliminary evidence suggests Alzheimer’s Disease (AD) risk may be related to impaired renal function but the association of variation in levels of commonly used AD biomarkers with kidney function are unknown.

**Objective:** To investigate the association between estimated glomerular filtration rate (eGFR), CSF levels of AD biomarkers: amyloid beta1–42 (Aβ42), Tau or phosphorylated Tau181 (pTau).

**Design, Setting, and Participants:** We conducted an analysis using data from participants enrolled in two research protocols at the Goizueta Alzheimer’s Disease Research Center that had simultaneous measurements of serum creatinine at the time of their Cerebrospinal fluid (CSF) collection (N=973). The participants had a mean age of 66.52 years, 23.33% were African American, and 63% were women, with 42.46% having mild cognitive impairment (MCI). The estimated glomerular filtration rate (eGFR) was obtained from chronic kidney disease Epidemiology Collaboration. All participants had similar CSF collection procedures. Aβ42, Tau or pTau were measured on the Luminex ALZBIO platform. General linear models and individual data were used to assess relationships between biomarkers and eGFR.

**Results:** Lower eGFR was associated with lower Aβ42/Tau ratio (slope= 0.033 units, p<0.0001) and Aβ42 (slope=0.75, p=0.002) and higher Tau (slope= -0.39, p<0.0001) and pTau (slope= -0.13, p=0.0002). Although these associations remained significant after adjusting for cognitive status, we observed interactions between MCI and eGFR. This interaction revealed that the impact of eGFR on AD biomarker levels was more robust in individuals with cognitive impairment (interaction MCI*GFR p-values were 0.005 for Ab42, 0.04 for tau and pTau, and 0.05 for the ratio).

**Conclusion:** We found a significant association between eGFR with CSF AD-biomarkers that may differ by cognitive status. This suggests that kidney function should be considered both in the context of interpreting AD biomarkers as well as exploring potential systemic factors that may increase risk of AD. Future longitudinal studies need to further explore the impact of kidney function on the pathogenesis of AD and related Biomarkers.

**KEY POINTS:** 

**Question:** Are Cerebrospinal Fluid (CSF)AD-biomarker measurements impacted by kidney function?

**Findings:** In this analysis of data from 973 individuals who had both cerebrospinal fluid (CSF) AD-biomarkers (Aβ42, Tau, and pTau181) and kidney function measurements, there were significant associations between estimated glomerular filtration rate (eGFR) and measures of CSF AD-biomarkers. These associations were more pronounced in those with cognitive impairment.

**Meaning:** Kidney function may have a significant impact on AD-biomarker measurements in the CSF, especially in those in the early symptomatic stages of AD.

## Introduction

Alzheimer’s Disease (AD) biomarkers such as amyloid beta1–42 (Aβ42), Tau, phosphorylated tau (pTau), and more specifically the Aβ42/Tau ratio play a crucial role in the diagnosis of Alzheimer’s Disease.^1^ These biomarkers are also commonly used in clinical trials for screening and enrollment and the use of a combination of these biomarkers has been found to yield improved case identifications.^2-5^ The variation in levels of these AD biomarkers by demographics and comorbidities is still being explored with recent evidence suggesting that consideration of these variations is important.^6-8^ Kidney function may impact levels of disease markers, especially on plasma protein-based markers.^9^ Whether there is a similar impact on AD-biomarkers commonly measured in cerebrospinal fluid (CSF) remains unclear. Preliminary evidence has suggested that impaired kidney function is associated with cognitive impairment based on neuropsychological assessments.^10-12^ A recent analysis of the Whitehall II prospective study suggested a connection between a lower estimated glomerular filtration rate (eGFR) and the risk of clinical dementia diagnosis.^13^ It has also been suggested that the risk of Alzheimer’s disease is related to impaired renal function and its impact on levels of AD biomarkers in the plasma.^14,15^ Since the CSF is a highly controlled microenvironment, studying the association of kidney function with CSF biomarkers offers great insight into the kidney-brain axis. ^12,16^

In this study, we aimed to investigate the association between estimated glomerular filtration rate (eGFR), as a measure of kidney function, and the levels of AD biomarkers in the CSF—specifically, the Aβ42/Tau ratio and individual other individual AD biomarkers: Amyloid beta 1-42 (Aβ42), Tau, and phosphorylated Tau181 (pTau)—in individuals with and without cognitive impairment. By analyzing the data from participants enrolled in two research protocols, we sought to determine whether eGFR is associated with alterations in the levels of AD biomarkers in the CSF. we also explored potential interactions between kidney function, AD biomarker levels, and mild cognitive impairment (MCI) – a common precursor to AD, which might provide critical insights into the impact of renal health on cognitive decline especially in individuals in the early symptomatic stages of AD.

## Methods

We analyzed data from participants in studies conducted within the Goizueta Alzheimer’s Disease Research Center (ADRC; N= 973, mean age=66.5, 52% mild cognitive impairment (MCI), 31% AA, 62% women) who had simultaneous measurement of their serum creatinine during their visit for CSF collection. Participants were enrolled in two observational protocols that were approved by the Emory IRB (Institutional Review Board) and each participant signed a written informed consent. All participants underwent identical lumbar puncture procedures, CSF analysis, and kidney function measurements: Brain Stress Hypertension and Aging program (B-SHARP: n= 375 and ADRC Clinical Research in Neurology (CRIN): n=598). B-SHARP sample participants were identified either through a referral from the Goizueta ADRC or through strategic community partnerships with grass-roots health education organizations, health fairs, advertisements, and mail-out announcements. An appropriate study informant, defined as an individual who has regular contact with the participant for at least once a week (in person or by telephone), was also identified. The potential study participants attended a screening visit, during which they underwent cognitive testing. A study physician performed a clinical evaluation and lumbar punctures (LP). CRIN is an ADRC-affiliated research protocol that is offered to individuals undergoing lumbar puncture as part of their evaluation at the Emory Cognitive Neurology clinic. LP procedures and preanalytical protocols are identical in all our ADRC-affiliated research and clinical protocols.

Demographics (age, sex, race, education), anthropometrics (weight and height), medical diagnosis, and income levels were collected at baseline by interview. Following a fast of no less than 6 hours, CSF samples were collected via lumbar puncture using 24G Sprotte atraumatic spinal needles. Samples were collected in sterile polypropylene tubes, separated into 0.5cc aliquots, and stored at −80 °C. CSF Biomarkers, Aβ1-42, tau, and Ptau181, were measured using the multiplex xMAP Luminex platform (Luminex Corp, Austin, TX) with Innogenetics (INNO-BIA AlzBio3; Ghent, Belgium; for research use– only reagents) immunoassay kit–based reagents. Estimated GFR was calculated using the Chronic Kidney Disease Epidemiology Collaboration (CKD-EPI) equation.^17^ CKD-EPI is the most widely used method to calculate eGFR which is calculated using this equation: 41 × min (Scr/κ, 1)α × max (Scr/κ, 1)- 1.209 × 0.993Age × 1.018 [if female] × 1.159 [if African American] (where Scr is serum creatinine in μmol/L, κ is 61.9 for females and 79.6 for males, α is -0.329 for females and -0.411 for males, min indicates the minimum of Scr/κ or 1, and max indicates the maximum of Scr/κ or 1).

### Statistical analysis

Statistical analysis was performed with SAS statistical software version 9.4 M6. The statistical significance was defined as p < 0.05. Although the 2 protocols had identical procedures for the biomarkers and kidney function, we conducted an individual participant data metanalysis with a General linear model and “protocol” as the random effect to assess relationships between biomarkers and eGFR using the combined data.^18^ In addition to the eGFR/biomarker measurement analysis, we conducted an interaction analysis to assess if the associations differed by cognitive status.

## Results

### Participant Characteristics

The Study’s sample included 973 participants, consisting of 613 women (63%) and 360 men (37%). The mean age was 66.52 (SD: 8.83). The participant distribution included 227 African American patients (23.33%) and 746 Caucasian patients (76.67%). Among the participants, 413 patients (42.45%) were diagnosed with Mild Cognitive Impairment (MCI). The mean eGFR was 78 (standard deviation: 16.70) mL/min/1.73 m^2^ with no significant difference between participants with Normal Cognition (NC) or participants with Mild Cognitive Impairment (MCI). As expected, we found that individuals within the MCI group exhibited a substantially lower average Aβ42 level, and higher average Tau and pTau levels. The key characteristics of the study participants are provided in Table 1.

**Table 1:** Descriptive characteristics of the pooled sample^a^.

| Characteristic |  | Combined sample | NC | MCI | P-value<sup>b</sup> |
| --- | --- | --- | --- | --- | --- |
| N |  | 973 | 560 (57.6%) | 413 (42.4%) |  |
| Age, years mean (SD) |  | 66.52 (8.83) | 64.7 ± 8.3 | 69.0 ± 8.9 | <0.001* |
| Sex (%) | Female | 613 (63%) | 383(68.4%) | 230(55.7%) | <0.001* |
|  | Male | 360 (37%) | 177(31.6%) | 183(44.3%) |  |
| Race (%) | AA | 227 (23.33) | 114(20.4%) | 113(27.4%) | 0.011* |
|  | White | 746 (76.67%) | 446(79.6%) | 300(72.6%) |  |
| eGFR mean (SD) |  | 78.00 (16.70) | 79.2 (15.9) | 76.3 (17.6) | 0.007* |
| Aβ42, pg/ml: mean (SD) |  | 385.15 (178.22) | 457.2 (184.1) | 287.5 (111) | <0.001* |
| Tau, pg/ml mean (SD) |  | 66.75 (43.29) | 51.3 (23.4) | 87.6 (53.9) | <0.001* |
| pTau pg/ml mean (SD) |  | 31.52 (22.08) | 24.9 (11.9) | 40.6 (28.6) | <0.001* |
| Aβ42/tau Ratio mean (SD) |  | 8.06 (6.09) | 10.7 (6.4) | 4.5 (3.1) | <0.001* |

<sup>a</sup> **Abbreviations:** - **NC:** Normal Cognition, **MCI:** Mild Cognitive Impairment, **SD:** standard deviation, **N:** Sample size, **eGFR:** Estimated Glomerular Filtration Rate, **Aβ42:** Amyloid-β42, **Tau:** Tau protein, **pTau:** Phosphorylated Tau protein, **AA:** African American.
<sup>b</sup> P-values obtained through a pooled analysis using t-test for continuous data, and chi-square for discrete data.

### Association of eGFR with CSF AD-biomarkers in Full sample

We observed a significant association between eGFR and the Aβ42/Tau ratio (0.033, p<0.0001). We also found significant associations between eGFR and all three CSF AD-biomarkers: Aβ42 (slope per unit increase: 0.75, p=0.002), Tau (-0.39, p<0.0001), and pTau (-0.13, p=0.0002). The results show that for every unit increase in eGFR, there is a corresponding change in CSF AD-biomarkers as follows: an increase in both the Aβ42/Tau ratio, and Aβ42 and a decrease in both Tau, and pTau. (Table 2) Furthermore, our analysis showed that each unit increase in eGFR corresponded to an approximate increase of 0.52 in Aβ42 levels, however, for Tau and pTau, we found that each unit increase in eGFR corresponded with a decrease in Tau and pTau levels in the CSF, even after adjusting for MCI status. (Table 2) Further, individuals within the MCI group exhibited a substantially lower average Aβ42 level measuring approximately (-88.88 pg/ml), lower Aβ42/Tau ratio (-2.85), and higher levels of Tau (40.51pg/ml), and pTau (17.03 pg/ml) after adjusting for eGFR. (Table S.2)

**Table 2:** Association of CSF biomarkers with estimated Glomerular filtration rate from the pooled sample^ab^.

| **Biomarker** | **Change<sup>c</sup> per unit of eGFR (95% CI)** | **P-value** | **MCI adjusted Change<sup>d</sup> per unit of eGFR (95% CI)** | **P-value** |
| --- | --- | --- | --- | --- |
| **Aβ42 pg/ml** | 0.75, (0.27,1.23) | 0.002 | 0.52 (0.03, 1.01) | 0.04 |
| **Tau pg/ml** | -0.39, (-0.55, -0.23) | <0.001 | -0.23 (-0.38, -0.09) | 0.001 |
| **pTau pg/ml** | -0.13, (-0.19, -0.06) | 0.0002 | -0.07 (-0.14, -0.01) | 0.02 |
| **Aβ42/Tau Ratio** | 0.033, (0.02,0.05) | <0.001 | 0.03 (0.01, 0.04) | 0.002 |

<sup>a</sup> **Abbreviations:** **CI 95%:** 95% Confidence Interval, **eGFR:** Estimated Glomerular Filtration Rate, **MCI:** Mild Cognitive Impairment, **Aβ42:** Amyloid- β42, **Tau:** Tau protein, **pTau:** Phosphorylated Tau protein.
<sup>b</sup> *P*-values were obtained through a meta-analysis using linear mixed-effects models and ‘Study’ as a random effect.
<sup>c</sup> Change is the Beta coefficient for the change in the CSF biomarker Levels per change in eGFR.
<sup>d</sup> Change is the Beta coefficient for the change in the CSF biomarker Levels per change in eGFR adjusted for MCI

### The impact of MCI on the association between CSF AD-biomarkers and eGFR

Our subsequent analysis aimed to explore the potential interaction among CSF AD biomarkers, eGFR, and MCI. We found a notable difference in the variation of CSF AD biomarkers with eGFR based on cognitive status, where the variation in AD biomarker levels was significant only in the MCI group.

However, in the NC group the variation with eGFR did not reach statistical significance. (Table 3) This observation indicates that the impact of eGFR on AD biomarker levels is more pronounced in individuals with cognitive impairment. (Figure 1) Finally, after adjusting for hypertension and diabetes, which was available only for a subsample (N=375, 50.74% with hypertension and 12.76% with diabetes), the association between the Aβ42/Tau ratio, Tau, and pTau, with eGFR remained significant except for Aβ42. (Table S.3)

**Table 3:** Interaction between MCI, CSF biomarkers, and eGFR from the pooled sample ^ab^.

| Biomarkers | Effects | $\beta$ | CI 95% | P-value<sup>c</sup> |
| --- | --- | --- | --- | --- |
| **Aβ42 pg/ml** | eGFR * MCI | 1.37 | (0.4, 2.33) | 0.005 |
| **Tau pg/ml** | eGFR * MCI | -0.30 | (-0.59, -0.01) | 0.04 |
| **pTau pg/ml** | eGFR * MCI | -0.13 | (-0.25, 0) | 0.05 |
| **Aβ42/Tau Ratio** | eGFR * MCI | 0.03 | (0, 0.06) | 0.05 |

<sup>a</sup> **Abbreviations:** $\beta$ : Beta coefficient for interaction between CSF biomarkers, **CI 95%:** 95% Confidence Interval, **MCI:** Mild Cognitive Impairment, **eGFR:** Estimated Glomerular Filtration Rate, **Aβ42:** Amyloid- β42, **Tau:** Tau protein, **pTau:** Phosphorylated Tau protein.
<sup>b</sup> Model includes the interaction term eGFR\*MCI and uses Study as a random effect.
<sup>c</sup> *P*-values obtained through Meta-analysis of the data from both studies (BSHARP & CRIN).

**Figure 1.**
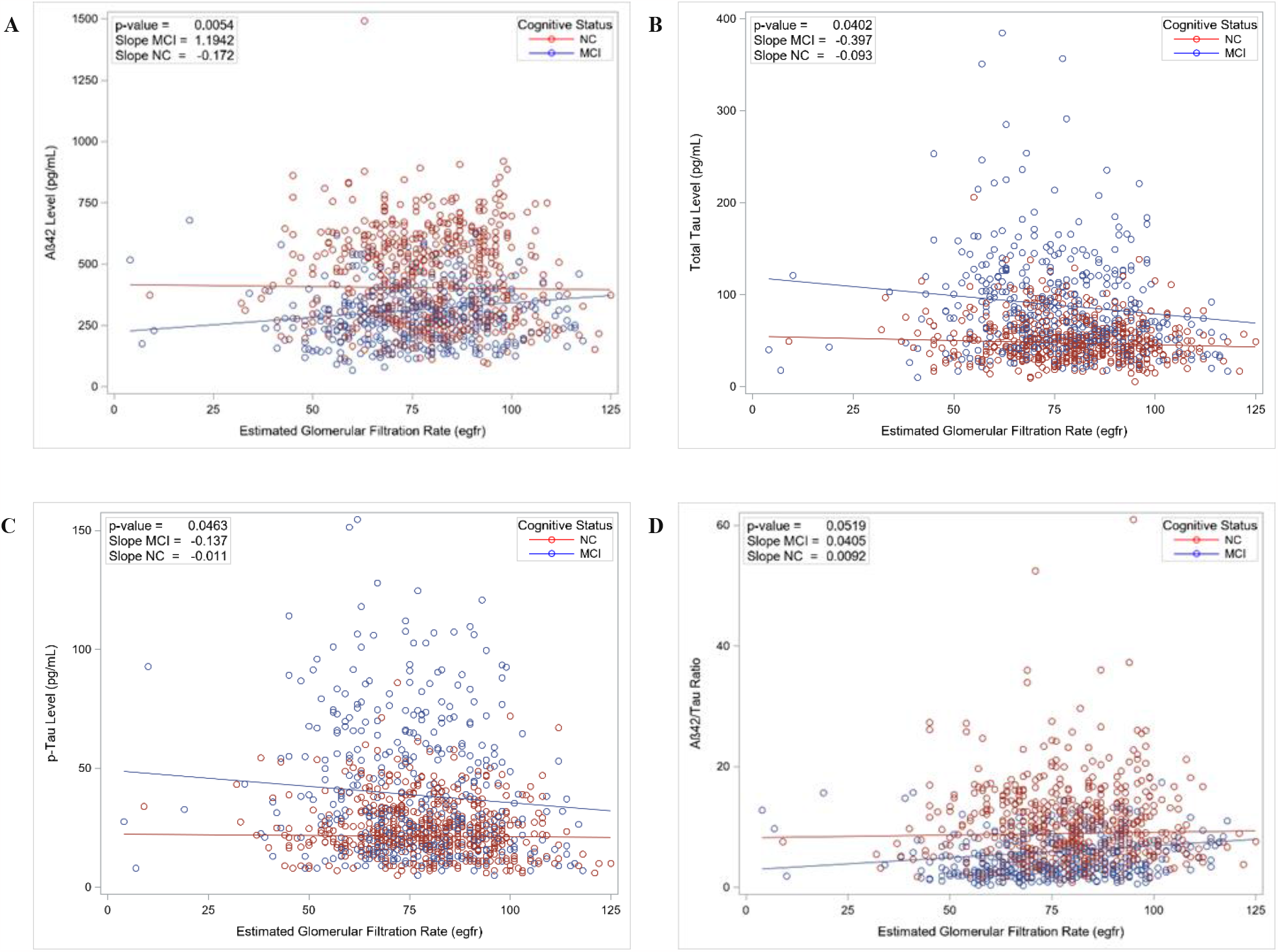
Impact of MCI on the Association Between CSF AD-Biomarker levels and eGFR. Interaction analysis performed using linear mixed models to assess differences in the relationship between cerebrospinal fluid (CSF) Alzheimer’s disease (AD) biomarker levels: (A) Aβ42, (B) Tau, (C) pTau, and (D) Aβ42/Tau Ratio and estimated glomerular filtration rate (eGFR) based on Cognitive status. The slope and intercept are plotted separately for individuals with Mild Cognitive Impairment (MCI) and those with Normal Cognition (NC). The p-value presented is for the interaction term (eGFR*MCI), it suggests a significant difference in the association between these variables based on cognitive status.

## Discussion

In our present study of a large cohort of older adults with CSF biomarker quantifications, we found kidney function was associated with the levels of AD biomarkers in the CSF. Although these associations remained significant after adjusting for cognitive status, we observed significant interactions between MCI and eGFR where the impact of this association is more pronounced in individuals with MCI. The significant association between CSF biomarkers and both eGFR and MCI, even after adjusting for confounders reinforces the notion from previous studies of a more direct influence of kidney health on AD-related pathology.^19^ Earlier studies have consistently indicated a link between kidney function, cognitive impairment, and dementia.^10-13^ Additionally recent studies have found that the kidneys play a physiological role in the clearance of Aβ from the blood and brain in both humans and animals,^20,21^ as well as Tau having a physiological role in the kidneys.^22^ Furthermore, studies have suggested that Aβ from the periphery might be contributing to AD pathogenesis by increasing the levels of Aβ crossing the blood brain barrier (BBB) and inducing Aβ-related pathologies in the brain.^23,24^ Therefore, given it has been established that lower Aβ42 levels in the CSF or plasma are associated with increased amyloid deposition in the brain by many clinical studies ^23,25-29^ and the amyloid hypothesis that suggests AD is due to the imbalance of Aβ,^30^ a possible explanation for our findings is the decline in kidney function resulted in decreased clearance and therefore accumulation of AD pathology^.31,32^ Another possible explanation is that the decline in kidney function contributed to an increase in production of AD biomarkers in the brain. In both scenarios, the association of kidney function with AD biomarkers needs further exploration. It is also worth noting that the associations between kidney function and Tau and Aβ42/tau ratio were significant after adjusting for hypertension and diabetes, suggesting that our findings are independent of common risk factors for kidney function loss. Our finding that adjusting for hypertension and diabetes renders the Aβ42-kidney function association not significant may imply that it is related to kidney disease risk factors. This supports previous studies that demonstrated an association between increased plasma levels of Aβ and vascular disease in both the brain and the periphery including hypertension, and diabetes.^24^ However, it is important to note that this analysis was only conducted on a subsample and hence need to be interpreted with caution.

This is to our knowledge one of the few first large studies that reported the association of kidney function with CSF AD biomarkers and its interaction with cognitive status. This study highlights the need to incorporate kidney function in studies of AD risk factors and the need to consider renal health when interpreting the results of AD biomarkers^33,34^ within clinical evaluations and trials particularly during the early symptomatic stages of AD. This also adds an additional factor to our understanding of the complex multifactorial processes contributing to AD and its biomarkers.

### Limitations

This study has a few limitations that are related to the cross-sectional nature of this analysis. Nevertheless, the observation of an association between kidney function and AD biomarkers cross-sectionally is significant. However, considering that the AD biomarkers were measured at a single point, whether kidney function would influence these measures in a longitudinal study remains an important and unexplored phenomenon.

### Future Directions

It is important that future studies investigate the association with other known comorbidities such as microvascular dysfunction, vitamin D metabolism, erythropoietin variation, and alterations in the renin-angiotensin system and their effects on AD biomarker variations. Studying their effects on AD biomarker variations could offer insights into the mechanisms for increased risk of AD in high-risk populations such as older adults, African Americans (AA) or those with Diabetes and hypertension.

## Conclusion

In conclusion, our study provided insights into the complex association between kidney function, AD biomarkers, and cognitive impairment. The findings suggest a significant association between kidney function and variation in levels of AD biomarkers in the CSF (Aβ42, tau, and pTau), which were stronger in individuals with MCI. This suggests that kidney function should be considered in the context of interpreting AD biomarkers in clinical evaluations and clinical trials, especially in those in the early symptomatic stage of AD. Future studies need to further explore the impact of kidney function and other comorbidities on the interpretation of AD Biomarkers.

## Data Availability

All data produced in the present study are available upon reasonable request to the corresponding authors

## Corresponding Author

Ihab Hajjar, MD, Department of Neurology, University of Texas Southwestern Medical Center, 5323 Harry Hines Blvd, 4^th^ floor, Dallas, TX 75390-8806, USA.

## Author Contributions

Dr Hajjar had full access to all of the data in the study and takes responsibility for the integrity of the data and the accuracy of the data analysis.

*Concept and design:* Hajjar.

*Acquisition, analysis, or interpretation of data:* Hajjar

*Drafting of the manuscript:* Hajjar, Neal, Yang.

*Critical revision of the manuscript for important intellectual content:* All authors.

*Statistical analysis:* Hajjar, Yang.

*Administrative, technical, or material support:* Hajjar

