## Supplemental Tables for "Alzheimer’s Disease cerebrospinal fluid Biomarkers and kidney function in normal and cognitively impaired older adults"

**Supplement:**

**Table S.1: Descriptive characteristics of the sample in the two protocols used and the combined sample.^[1]^**

| **Characteristic** | | **B-SHARP** | **CRIN** |
| --- | --- | --- | --- |
| **N** | | 375 | 598 |
| **Age** | | 65.56 (7.95) | 67.37 (9.24) |
| **Sex (%)** | **Female** | 377 (61.10%) | 378 (63.21%) |
|  | **Male** | 240 (28.90%) | 220 (36.79%) |
| **Race (%)** | **African Americans** | 148 (39.47%) | 79 (13.21%) |
|  | **White** | 227 (60.53%) | 519 (86.79%) |
| **Mild Cognitive impairment** | | 214 (57.07%) | 199 (33.28%) |

**Table S.2: Meta-analysis with both eGFR and MCI in the model, no interaction terms^[2]^**

| **Variable** | **Effect** | **β** | **CI 95%** | ***P* value^[3]^** |
| --- | --- | --- | --- | --- |
| **Aβ42 pg/ml** | eGFR ml/min/1.73m2 | 0.52 | (0.03, 1.01) | 0.04 |
|  | MCI | -88.88 | (-105.77, -71.99) | <0.001 |
| **Tau** **pg/ml** | eGFR ml/min/1.73m2 | -0.23 | (-0.38, -0.09) | 0.002 |
|  | MCI | 40.51 | (35.46, 45.56) | <0.001 |
| **pTau** **pg/ml** | eGFR ml/min/1.73m2 | -0.07 | (-0.14, -0.01) | 0.02 |
|  | MCI | 17.03 | (14.87, 19.18) | <0.001 |
| **Aβ42/Tau Ratio** | eGFR ml/min/1.73m2 | 0.03 | (0.01, 0.04) | 0.002 |
|  | MCI | -2.85 | (-3.4, -2.3) | <0.001 |

**Table S.3: adjusted analysis for Hypertension and Diabetes (For** **BSHARP Only) ^[4]^**

| **Variables** | | **β** | ***P*-value^[5]^** |
| --- | --- | --- | --- |
| **Aβ42** | **eGFR** | 0.40 | 0.15 |
|  | **MCI** | -20.51 | 0.04 |
|  | **HTN** | -25.63 | 0.01 |
|  | **DM** | -31.38 | 0.04 |
| **Tau** | **eGFR** | -0.44 | <0.001 |
|  | **MCI** | 15.65 | <0.001 |
|  | **HTN** | 14.75 | 0.001 |
|  | **DM** | 5.33 | 0.42 |
| **pTau** | **eGFR** | -0.11 | 0.01 |
|  | **MCI** | 6.99 | <0.001 |
|  | **HTN** | 3.91 | 0.01 |
|  | **DM** | 2.15 | 0.35 |
| **Aβ42/tau Ratio** | **eGFR** | 0.03 | 0.003 |
|  | **MCI** | -0.87 | 0.007 |
|  | **HTN** | -1.35 | <0.001 |
|  | **DM** | -0.35 | 0.47 |

1. **Abbreviations**: - **N:** Sample size, **SD:** standard deviation, **B-SHARP:** Brain Stress Hypertension and Aging program, **CRIN:** ADRC Clinical Research in Neurology.
2. **Abbreviations**: **β:** Beta coefficient**, CI 95%:** 95% Confidence Interval **eGFR**: Estimated Glomerular Filtration Rate, **MCI:** Mild Cognitive Impairment, **Aβ42:** Amyloid-β42, **Tau**: Tau protein, **pTau:** Phosphorylated Tau protein,
3. *P*-values obtained through Meta-analysis using linear mixed-effects models and ‘Study’ as random effect. Model includes (eGFR, and MCI).
4. **Abbreviations**: **B-SHARP:** Brain Stress Hypertension and Aging program, **β:** Beta coefficient**, eGFR**: Estimated Glomerular Filtration Rate, **MCI:** Mild Cognitive Impairment, **HTN:** Hypertension, **DM:** Diabetes Miletus, **Aβ42:** Amyloid-β42, Tau: Tau protein, pTau: Phosphorylated Tau protein.
5. *P*-values obtained through General linear model.
